# Multivariate association with disease progression and walking kinematics in relapsing-remitting multiple sclerosis

**DOI:** 10.1101/2025.05.07.25327172

**Authors:** Sumire D. Sato, Sutton B. Richmond, Clayton W. Swanson, Kristin A. Johnson, Brett W. Fling, Rachael D. Seidler

## Abstract

**Highlights:**

Multiple sclerosis disease duration was not associated with gait speed
Multivariate models with gait kinematics are associated with disease duration
Disease duration is linked to step width variability and ankle pitch at toe-off

Declines in walking function commonly occur in people with multiple sclerosis (MS). Walking is a continuous movement and a complex motor task that requires precise timing and scaling of activation across many muscles. The objectives of this study were to identify kinematic gait characteristics during a 2-minute walk test that are associated with disease duration in individuals with relapsing-remitting MS (n=45). Participants were instrumented with inertial measurement units from APDM (APDM Inc, Portland, OR, USA) and performed the 2-minute walk test twice: once at their self-selected speed and another at their fastest, safe speed. Gait speed was not associated with MS disease duration (Self-selected: p= 0.180; Fast: p=0.167). Canonical correlation analysis showed that disease progression was strongly associated with lateral step width variability and ankle pitch at toe-off, which was also reflected in the multiple linear regression models. Both self-selected and fast-speed kinematics were associated with disease duration (Self-selected: p=0.007; Fast: p=0.004). Thus, we identified a multivariate model that is associated with disease duration in highly ambulatory MS individuals. Our study highlights the importance of considering individual differences in specific gait kinematics (such as lateral step width variability and ankle pitch at toe-off) when assessing disease progression in MS. This approach may provide more personalized insights into the impact of MS on mobility and help tailor interventions to improve gait and overall function in affected individuals. Future research should continue exploring these kinematic parameters to better understand their role in MS progression and develop targeted therapeutic strategies.

## INTRODUCTION

Multiple sclerosis (MS) is an immune-mediated inflammatory demyelinating disease of the central nervous system, which involves damage to the myelin sheath that insulates nerve fibers. 85-90% of patients with MS follow a relapsing-remitting course at disease onset, characterized by episodes of new or worsening symptoms (relapses) followed by partial or complete recovery (remissions)[1]. Disease progression is typically quantified with the Expanded Disability Status Scale (EDSS), neuroimaging, clinical assessments, and patient-reported outcomes[2–4]. Walking function is a crucial construct in many outcome measures to assess disability in people with MS[5]. Walking assessments, such as the Timed 25-foot Walk Test (T25FW), are commonly used to evaluate gait and mobility[6]. Longer tests, like the 2-minute walk test, can also assess gait characteristics affected by fatigue – a major complaint in people with MS[7].

However, walking assessments often fail to detect subtle or early-stage sensorimotor dysfunction in MS, particularly in those with low to moderate EDSS scores[8].Understanding the variability in disease progression at the early stages of MS is crucial for developing personal treatment plans and improving patient outcomes.

Here, our main interest was to examine different spatiotemporal characteristics of walking. Walking is a continuous movement and a complex motor task that requires precise timing and scaling of muscle activation across many muscles. Many kinematic gait characteristics significantly differ between people with MS and healthy populations [9–12]. However, there is considerable inter- and intra-individual variability in gait characteristics. Even in healthy adults, there are distinct individual characteristics in walking, influenced by various factors such as biomechanics, neuromuscular control, and personal habits[13, 14]. This individuality in walking patterns underscores the complexity of gait analysis and the need for personalized approaches in studying and addressing gait abnormalities.

Changes in different spatiotemporal characteristics of walking may identify disease progression of MS. For example, higher cadence is associated with lower risk of mortality, higher cardiovascular fitness, and decreased fall risk[15, 16]. An increase in double support time may enhance stability in those with balance difficulties [17]. Longer stride length may be a strategy to improve stability but it also indicates stronger and more flexible leg musculature[18]. Lateral step width variability is a measure of deviations in the width of the steps taken; an increase in lateral step width variability may indicate difficulty with precision movement planning and execution[19–21]. Higher foot clearance may indicate that an individual has better neuromuscular control and will reduce the risk of tripping and falling[22]. Ankle pitch at toe-off and heel-strike could give information about effective push-off and landing strategies, and a decrease in these angles may indicate increased spasticity or stiffness in the lower limb[23]. Many studies have focused on examining the influence of single gait variables as risk factors or predictors of diseases that impact gait[24]. A multifactorial model of gait characteristics in multiple sclerosis demonstrated significant kinematic differences between MS severity levels [25]. However, the extent to which gait characteristics can adequately represent the disease progression in MS remains unclear. Multivariate models considering different gait characteristics may provide important insights into MS disease progression and individualized rehabilitation strategies.

Here, we focus on the relapsing-remitting subtype of MS (RRMS), which shows variability in patient outcomes but patients are highly ambulatory (i.e., independent). The objectives of this study were to identify gait characteristics that are associated with disease duration in individuals with RRMS. We examined gait characteristics at participants’ ‘fastest safe’ speed and ‘self-selected’ speed; the fastest gait speed may provide information about the individual’s highest capacity/movement ability, while self-selected speed is a safer way to test walking in the clinic. We hypothesized that (1) gait speed would not be associated with MS disease duration, (2) fast speed gait kinematics would predict disease duration better than self-selected speed kinematics, and (3) we explored multivariate kinematic models that predicted associations with MS disease duration.

## METHODS

### Participants

45 people with RRMS were included in this study. Participant characteristics are presented in Table 1. Participants were combined from two different studies that followed the same exclusion/inclusion criteria[10, 26]. All participants had a confirmed diagnosis of RRMS by a clinician. Participants were excluded from this study if they were unable to stand unassisted on a firm surface, scored > 4.0 on the self-administered EDSS (indicating inability to ambulate independently), had a joint replacement, musculoskeletal/vestibular dysfunction, or any other neurological impairment other than MS. All participants gave written informed consent before the studies in accordance with the Declaration of Helsinki and with the protocols approved by the Institutional Review Board of Colorado State University (#1664).

**Table 1.**
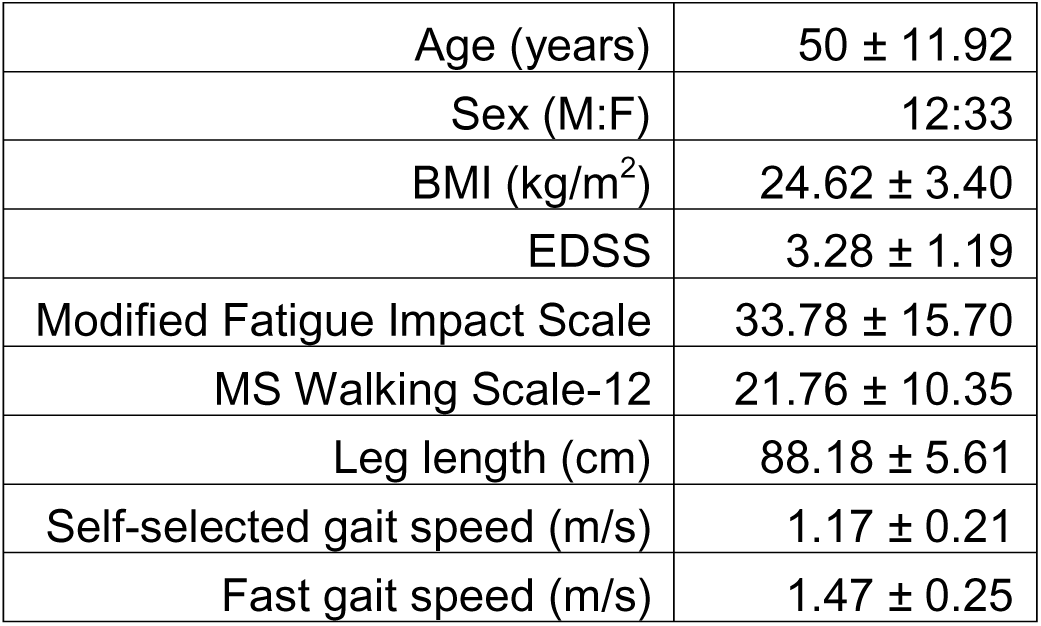
Participant characteristics. EDSS = Expanded Disability Status Scale (Score range: 0-10; Higher score indicates more disability). Modified fatigue impact scale score range: 0-84; Higher score indicates greater impact of fatigue on a person’s activities. MS Walking Scale-12 = 12-item Multiple Sclerosis Walking Scale (Score range: 0-60; Higher scores indicate greater impact on walking ability).

### Walking assessment

Participants were instrumented with six, tri-axial inertial measurement units from APDM (Version 2.0, APDM Inc, Portland, OR, USA), placed on the dorsum of each foot, on the dorsum of each wrist, at the lumbar level (L4-L5), and the sternum (Figure 1A).

**Figure 1.**
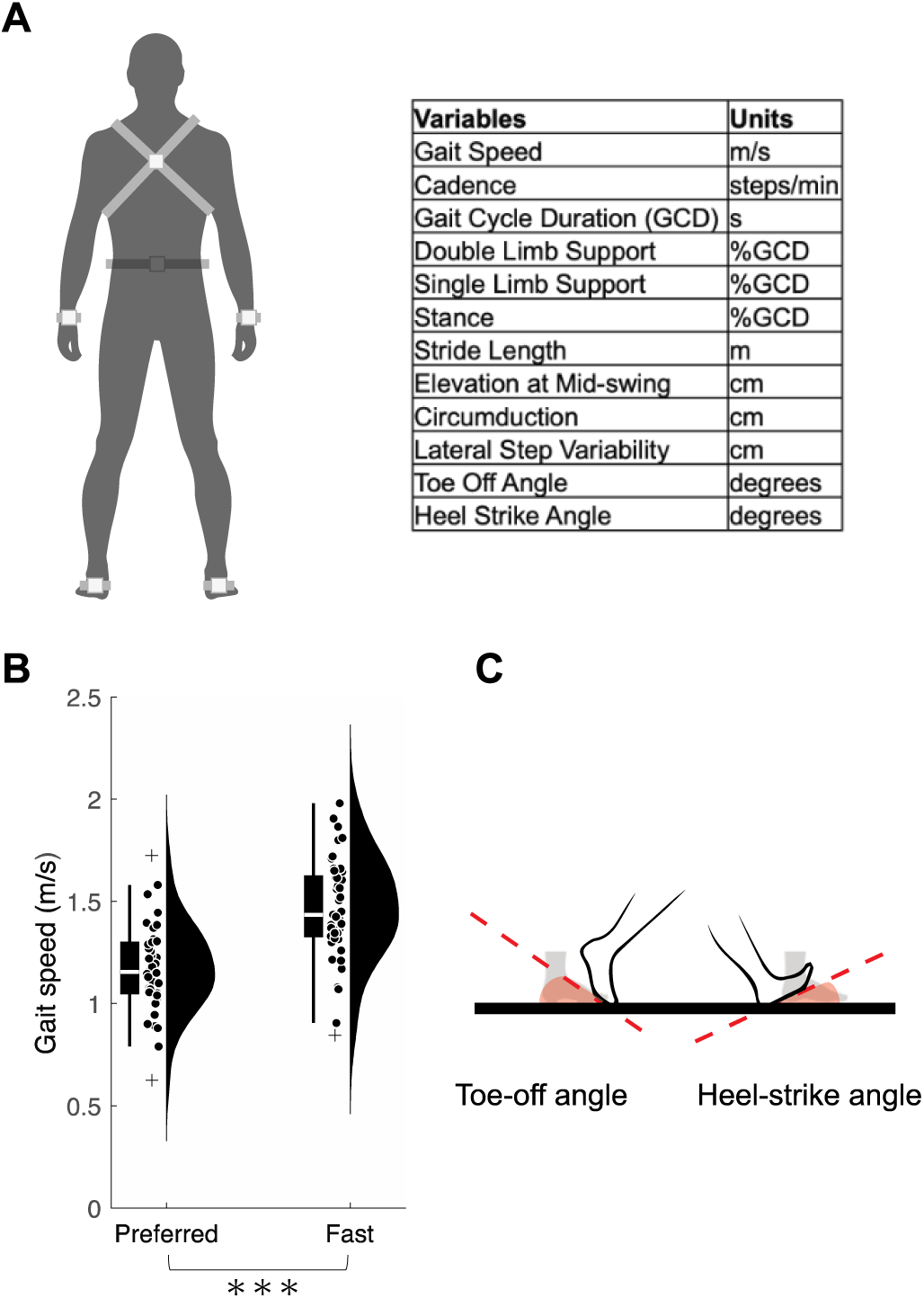
Experimental methods. A. Inertial sensor placement. Participants were instrumented with six tri-axial inertial monitoring units from APDM (Version 2.0, APDM Inc, Portland, OR, USA), placed on the dorsum of each foot, on the dorsum of each wrist, at the lumbar level (L4-L5), and the sternum. B. Spatiotemporal variables and their units. Descriptions of each variable are in Supplementary Table 1. C. Gait speed of the self-selected 2MWT and fast 2MWT. Speeds between the two trials were significantly different (p < 0.001). C. Depiction of toe-off angle and heel-strike angle. Note these are ankle *pitch* (angle of the foot compared to the floor), not the ankle angle.

**Figure 2.**
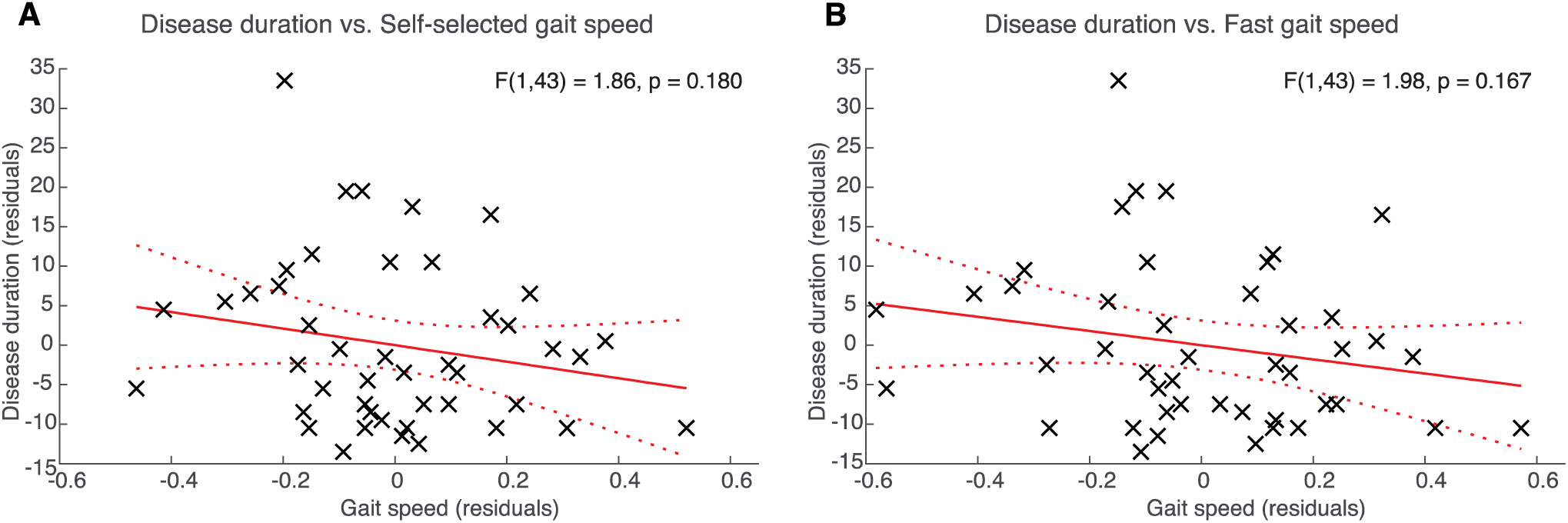
Partial correlations between disease duration and gait speed during preferred speed 2MWT (A) and fast speed 2MWT(B). Solid red line = best-fit line. Dotted red lines = 95% confidence bounds.

All inertial measurement units were sampled at 128 Hz (Morris et al., 2019). Participants underwent two, 2-minute walking tests (2MWT), where they walked continuously (*back and forth*) at a “self-selected pace” down a 110-foot hallway for the initial gait trial, and then at a “fast-pace” during the second trial. Each trial was performed without an assistive device and free of distractions or obstacles. For the self-selected walking trial, participants were asked to walk comfortably as if “casually navigating a grocery store or walking around their home”. For the fast walk, all participants were asked to walk “as fast as safely possible, without breaking into a jog or run.” Gait speed was significantly faster for the ‘fast’ trial than for the ‘self-selected’ trial (Paired t-test: t(44) = 16.02, p < 0.001; Figure 1B).

We quantified gait characteristics for each trial using the clinical user interface of Mobility Lab v2 (APDM Inc, Portland, OR, USA). The spatiotemporal parameters included here were gait speed, cadence, double support time (% gait cycle), stride length, foot clearance, lateral step width variability, circumduction, ankle pitch at heel-strike, and ankle pitch at toe-off (for descriptions of the spatiotemporal parameters, see Supplementary Table 1). All spatiotemporal parameters were calculated from stride pairs established during segments of straight-line walking only[27]; any deceleration or acceleration phases of turn negotiation (entering, during, and exiting) were excised from the analysis via the automated algorithms-this method has been tested for validity and repeatability[28–30].

### Statistical analysis

To assess the association between disease duration and walking speed, we used a partial correlation corrected for sex. This adjustment is necessary as men often experience a more aggressive MS disease course compared to women[31].

The relationship between disease duration and kinematics during self-selected and fast-speed 2MWT was assessed with a canonical correlation analysis. Canonical correlation analysis is a multivariate analytical approach that measures the relationship between two datasets. We identified a ‘Disease progression dataset’ with gait speed and EDSS score. We determined multi-collinearity with a Pearson’s correlation analysis. for the ‘Kinematics dataset.’ We used r = 0.8 for the cut-off for multicollinearity [32]. We double-checked multicollinearity by calculating the variance inflation factor (VIF). The final variables in the ‘Kinematics dataset’ included cadence, double support time, stride length, foot clearance, lateral step variability, circumduction, ankle pitch at heel-strike, and ankle pitch at toe-off. Canonical correlation analysis was conducted for each self-selected and fast-speed 2MWT kinematic data sets. All variables were standardized before canonical correlation analysis to make the canonical coefficients comparable across the variables. Canonical correlations range from 0 to 1, 0 being no relationship and 1 being a perfect relationship between the two datasets. As a post-hoc analysis, we tested backward stepwise multiple linear regression models with disease duration and the kinematic variables. Disease duration was corrected for sex by residuals obtained from a linear regression model.

All statistical significance was established with an alpha level = 0.05. We used RStudio (R version 4.3.0) for analysis. For canonical correlation analysis results, we applied the “CCA” package[33].

## RESULTS

### Associations between gait speed and disease duration

We first assessed whether gait speed is associated with disease duration. Neither self-selected (F(1,43) = 1.86, p = 0.180, r^2^ = 0.04) nor fast 2MWT gait speeds were associated with disease duration, corrected for biological sex (F(1,43) = 1.98, p = 0.170, r^2^ = 0.04).

### Canonical correlation analysis with kinematics during walking with self-selected speed

As gait speed was not correlated with disease duration, we assessed a multivariate model without gait speed. Double support time was highly correlated with single limb stance (r = −1.00), and stance time (r = 1.00); therefore, single limb stance and stance time were excluded from our canonical correlation analysis. VIF for each remaining predictor was < 6 (Table 2), suggesting multicollinearity is not a concern for our model.

**Table 2.** Variance inflation factor (VIF) for predictor variables for models with self-selected speed 2-minute walk test (2MWT) and fast speed 2MWT.

| Variable | VIF |  |
| --- | --- | --- |
|  | Self-selected | Fast |
| Cadence | 2.31 | 2.03 |
| Double support | 3.23 | 2.50 |
| Stride length | 5.94 | 4.90 |
| Foot clearance | 1.15 | 1.29 |
| Lateral step variability | 1.32 | 1.47 |
| Circumduction | 1.46 | 1.28 |
| Ankle pitch at HS | 3.15 | 3.04 |
| Ankle pitch at TO | 1.94 | 1.88 |

For self-selected speed 2MWT, only the first canonical correlation was statistically significant (First dimension: r =0.68, Wilk’s lambda (Λ) = 0.48, F(16,70) = 1.95, p = 0.03; Second dimension: r = 0.33, Λ = 0.89, F(7,36) =0.62, p =0.74). Figure 4 presents the standardized canonical coefficients for the first dimension. In the ‘Disease progression variates’, the canonical correlation was equally influenced by EDSS (Coef. = 0.79) and disease duration (Coef. = 0.52). For the ‘Kinematics variates’, the canonical correlation was most strongly influenced by ankle pitch at toe-off (Coef.=-0.73), ankle pitch at heel strike (Coef. = 0.61), and lateral step width variability (Coef.= 0.59). This observation demonstrates that individuals with RRMS with higher EDSS scores and longer disease duration have smaller ankle pitch at toe-off and larger ankle pitch at heel-strike and lateral step width variability.

**Figure 3.**
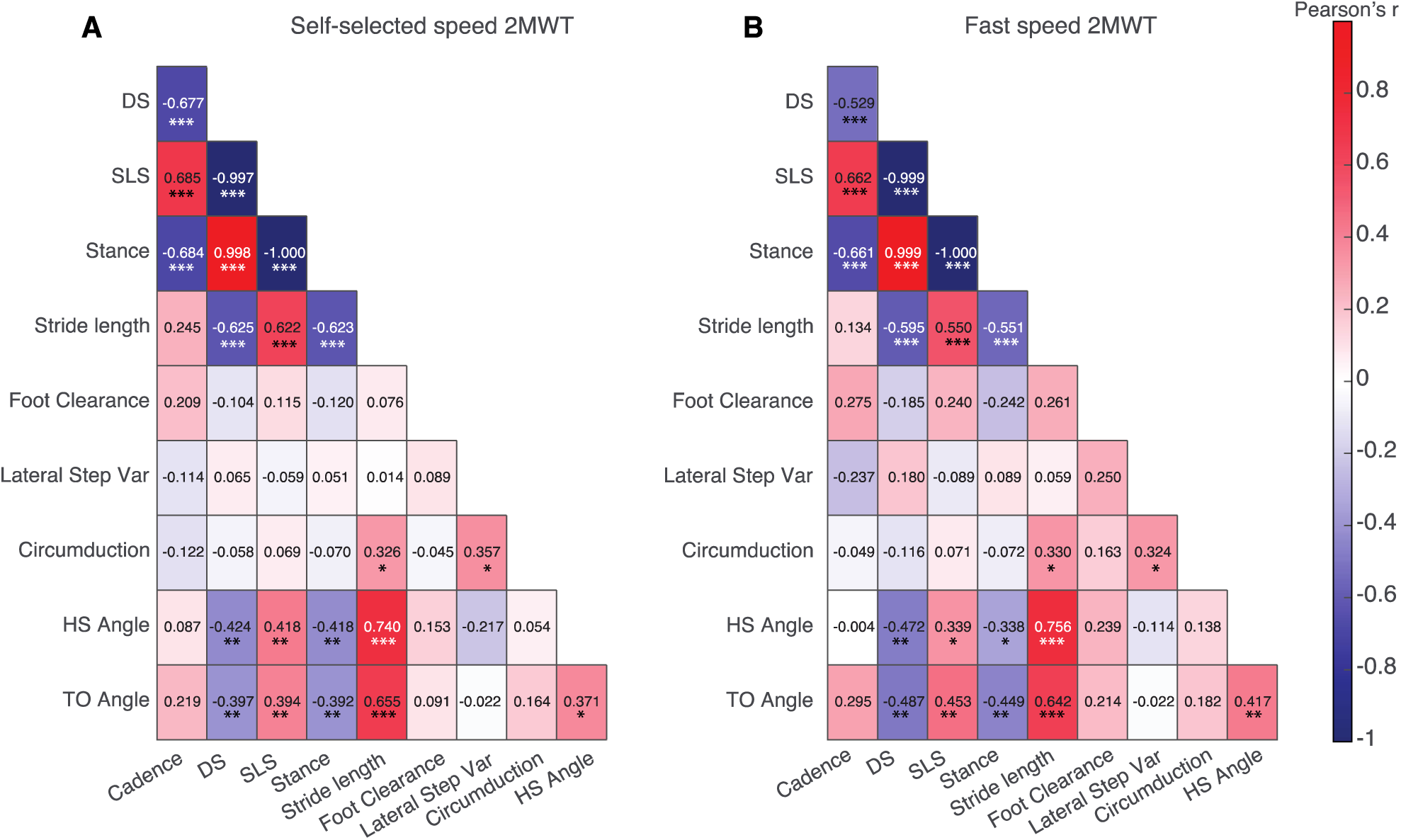
Pearson’s correlations between the kinematics variables. DS = Double support time. SLS = Single limb support time. HS Angle = Ankle pitch at heel-strike. TO Angle = Ankle pitch at toe-off. * = p-value 0.010 ≤ p < 0.050. ** = p-value 0.001 ≤ p < 0.009. ***= p < 0.001.

**Figure 4.**
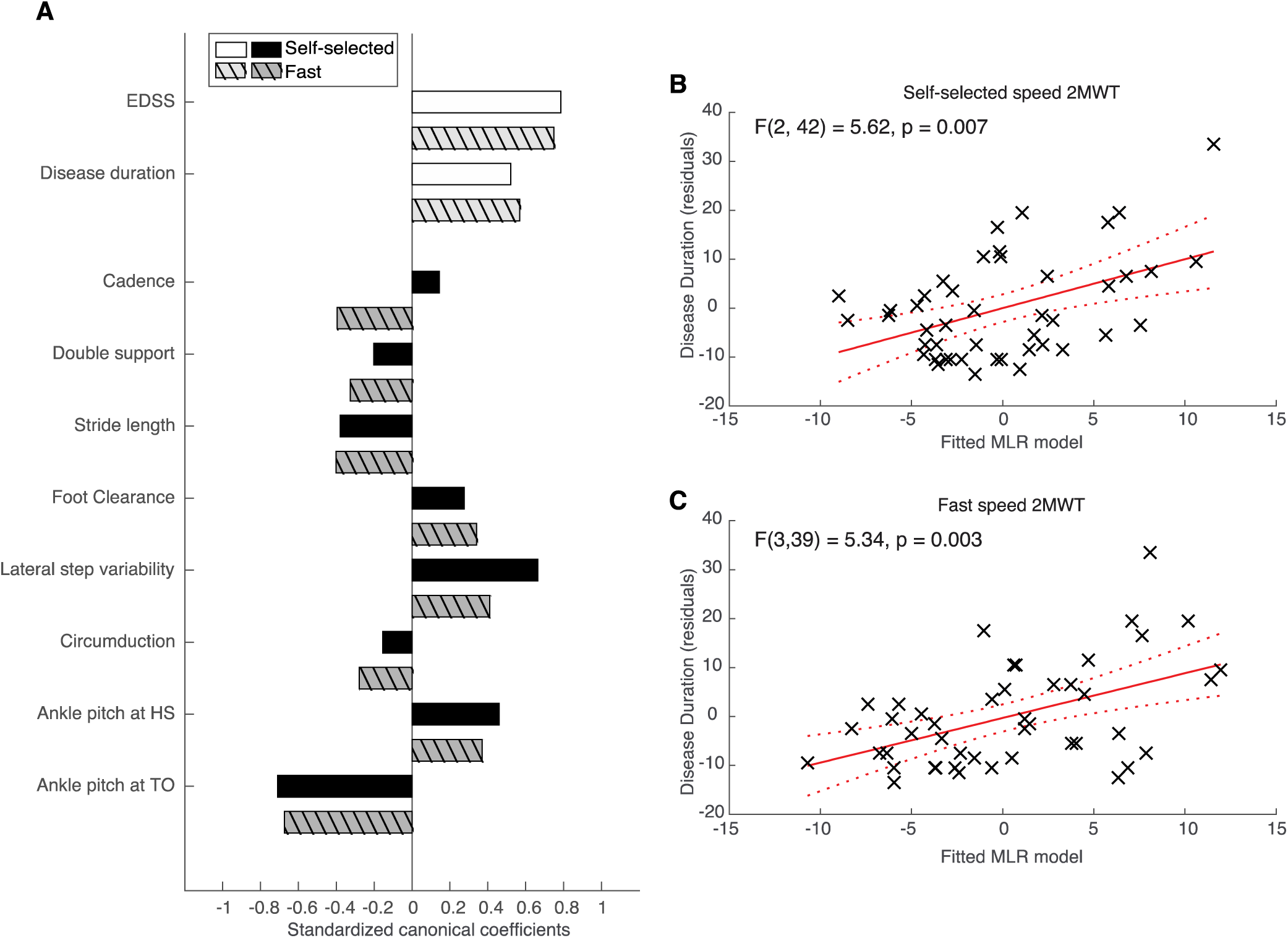
Canonical correlation analysis and multiple linear regression analysis. A. Standardized correlation coefficients. Only the first dimension showed a significant association of variance between the two datasets for both the preferred and fast 2MWT, which is presented here. White and light gray bars = Coefficients for the ‘Disease progression dataset’; Black and dark gray bars = Coefficients for the ‘Biomechanics dataset’. Solid bars = Coefficients for preferred speed 2MWT. Striped bars = Coefficients for fast speed 2MWT. B-C. Multiple linear regression models. Solid red line = best-fit line. Dotted red lines = 95% confidence bounds.

### Canonical correlation analysis with fast gait speed for 2MWT kinematics

The correlation table with the fast speed 2MWT kinematic variables showed that double support time was highly correlated with both single limb stance time (r = - 1.00),and stance time (r = 1.00); thus, these two variables were excluded from our canonical correlation analysis. VIF for each remaining predictor was < 5 (Table 2), suggesting multicollinearity is not a concern for our model.

Similar to the self-selected speed walking canonical correlation analysis, only the first dimension was statistically significant for fast speed (First dimension: r =0.71, Λ = 0.42, F(16,70) = 2.39, p = 0.007; Second dimension: r =0.39, Λ = 0.85, F(7,36) =0.91, p =0.512). Figure 4 presents the standardized canonical coefficients for the first dimension. In the ‘Disease progression variates’, the canonical correlation was influenced by EDSS (Coef. =0.75) and disease duration (Coef. =0.57). For the ‘Kinematics variates’, the canonical correlation was most strongly influenced by ankle pitch at toe-off (Coef.= −0.67), lateral step width variability (Coef.= 0.41), and stride length (Coef.=-0.40). This observation demonstrates that individuals with RRMS with higher EDSS scores, and longer disease duration have smaller ankle pitch at toe-off, larger lateral step width variability, and smaller stride length.

### Multiple linear regression with disease duration

The final model with self-selected speed kinematics predicting disease duration (controlled for sex) included lateral step width variability and ankle pitch at toe-off (F(2,42) = 5.62, p = 0.007). Lateral step width variability (B =4.58, β =0.31, p = 0.029) and ankle pitch at toe-off (B =-0.61, β =-0.33, p =0.020) were significant predictors, with the model explaining 21.1% of the variance (adjusted R² =0.173).

The final model with fast-speed kinematics predicting disease duration (controlled for sex) included lateral step width variability, ankle pitch at toe-off, and foot clearance (F(3,39) = 5.34, p = 0.004). Lateral step width variability (B = 4.66, β = 0.28, p = 0.049) and ankle pitch at toe-off (B = −0.70, β = −0.39, p = 0.008) were significant predictors. Foot clearance contributed to the model (B = 4.83, β = 0.25, p = 0.082), with the model explaining 29.1% of the variance (adjusted R² = 0.237).

## DISCUSSION

We found that gait speed was not associated with disease duration for individuals with RRMS, but we identified self-selected speed and fast speed walking kinematic variables associated with disease progression. A canonical correlation analysis showed that disease progression was largely influenced by lateral step width variability and ankle pitch at toe-off, which was also reflected in the multiple linear regression models. Both self-selected and fast speed kinematics were associated with disease duration.

### Gait speed is not associated with RRMS disease duration

Similar to previous studies that found mobility-based measures have limited ability to predict disease duration[34], we found that MS disease duration is not associated with walking speed. Our findings demonstrate that MS follows a complicated disease course that cannot be quantified by gait speed alone. Our study supports current clinical practice of considering multiple factors, including imaging, spasticity, cognitive function, and fatigue, to assess the progression of MS over time[2–4, 35]. In addition, the advancement of inertial measurement units or pose estimation software coupled with basic camera allows increased accessibility for spatiotemporal characteristics to be quantified in the clinic [36, 37]. Gait kinematics may provide additional insight into disease progression in MS.

### Disease duration is associated with lateral step width variability and ankle pitch at toe-off

In our canonical correlation model, those with longer disease duration and higher EDSS had larger lateral step width variability and smaller ankle pitch at toe-off. Several studies have identified greater intra-individual variability in gait kinematics in people with MS compared to healthy individuals[11, 12, 38]. Gait variability may be due to challenges during movement planning and execution. Motor variability is suggested to be due to stochastic neural processing or ‘noise’, which introduces randomness into motor commands[39]. The premotor and primary motor areas can reduce this variability through ‘quenching,’ where neural responses become more consistent when stimuli that drive the neurons are presented[19–21]. This reduction in neural variability helps increase the precision of movements, including lateral steps. Larger lateral step width variability indicates that those with longer MS disease duration are more likely to have difficulties with the ‘quenching’ process to maintain a steady and balanced walking pattern, which can be attributed to the progressive loss of motor control and coordination associated with MS.

Cerebellar control also plays a role in lateral step variability [40, 41]. The cerebellum coordinates and fine-tunes movements, including gait. When the cerebellum is damaged or impaired, it can lead to increased variability in the step width, and increased lateral step width variability is a characteristic feature of cerebellar ataxia[40, 41]. Recent work in MS has shown that the disease affects the cerebellum[42, 43]. Our models demonstrating increased lateral step width variability associated with longer disease duration may suggest that those who have had MS for a longer period may have more lesions in the cerebellum, presenting as increased lateral step width variability in their gait.

Ankle pitch at toe-off involves coordinated efforts between the central and peripheral nervous systems. It is driven by the strength of lower limb muscles to ensure efficient propulsion at toe-off. Interestingly, a study on gait kinematics in daily living found that ankle pitch at toe-off best discriminated individuals with mild-moderate MS from healthy controls[44]. This may indicate that people with longer disease duration have weaker peripheral limb musculature. Biofeedback has shown to improve ankle propulsion in older adults[45]; therapeutic interventions targeting specific ankle kinematics may help walking in those with MS. Whether the association between ankle-pitch at toe-off and disease duration is specific to MS is unclear and outside the scope of this study. However, our findings suggest that this kinematic measure may be important to quantify MS disease progression.

### Self-selected gait speed kinematics predict disease similar to fast gait kinematics

We examined gait characteristics at both ‘fastest safe’ and ‘self-selected’ speeds; we predicted that the models with gait kinematics from the fast walking trial would better predict disease duration than the self-selected speed trial. However, our results showed that the two multiple linear regression model outputs were similar in the variance explained. In the clinic, it is more common to assess 2MWT performance at a fast speed [46], but results from this study suggest that performing this test at a slower self-selected speed is just as informative. Self-selected speed is highly relevant to daily activities, and is less likely to be affected due to compensatory mechanisms[47]. In our study, our cohort was high-functioning (independent ambulation; also see discussion in *Limitations* section), so there may not have been compensatory mechanisms at the fast trial. Future studies should investigate gait characteristics both at self-selected and fast speed walking longitudinally with a larger cohort; self-selected speed gait analysis may provide additional valuable insights into a patient’s functional capabilities.

### Limitations

As mentioned in our methods section, the cohort analyzed here is from two distinct studies that were collected at different times. The two datasets were collected at the same location with the same procedures. The exception is that one cohort of participants wore shoes[26], while in the other they did not[10]. To ensure the robustness of our findings, we included the study location as a covariate in our models; this factor did not significantly contribute to the multiple linear regression model. Our results highlight the consistency of our findings across cohorts, reinforcing the reliability of our conclusions.

Another limitation of this study is that we focused on people with MS with independent ambulation. This represents a relatively high-functioning MS group and may limit the generalizability of our findings to a broader MS population. However, RRMS is the most commonly diagnosed form of MS[1]. By characterizing this subgroup’s gait and mobility patterns, our study provides valuable insights into the motor function and challenges faced by individuals with RRMS. Additionally, understanding the gait characteristics of higher-functioning individuals with MS can be informative for clinicians and researchers, especially when considering the progression of the disease and its impact on mobility over time.

### Conclusions

We found that longer MS disease duration was associated with increased lateral step width variability and ankle angle at toe-off in individuals with RRMS. MS is known for its heterogeneous presentation, but our study provides insight into neural mechanisms that may surface to a greater extent later in the disease course. This research highlights the importance of considering individual differences in gait when evaluating disease progression in MS. Future studies with larger cohorts may provide more personalized insights into the impact of MS on mobility and help tailor interventions to improve gait and overall function in individuals with MS.

## Data Availability

Data will be made available on resonable request.

