## Supplementary material for "Multivariate association with disease progression and walking kinematics in relapsing-remitting multiple sclerosis"

Supplementary Table 1. Gait kinematic variable definitions. (Modified from <https://www.apdm.com/wp-content/uploads/2015/05/02-Mobility-Lab-Whitepaper.pdf> )

| **Variables** | **Units** | **Definition** |
| --- | --- | --- |
| Gait Speed | m/s | The forward speed of the subject, measured as the forward distance traveled during the gait cycle divided by the gait cycle duration |
| Cadence | steps/min | The number of steps per minute, counting steps made by both feet |
| Gait Cycle Duration | s | The duration of a complete gait cycle measured from the left foot’s initial contact to the next initial contact of the left foot |
| Double Limb Support | %GCT | The percentage of the gait cycle in which both feet are on the ground |
| Single Limb Support | %GCT | The percentage of the gait cycle in which one foot is on the ground |
| Stance | %GCT | The percentage of the gait cycle in which the foot is on the ground |
| Stride Length | m | The forward distance traveled by the foot during a gait cycle |
| Elevation at Mid-swing | cm | The height of the foot sensor measured at mid-swing, relative to its start position while standing |
| Circumduction | cm | The maximum amount that the foot travels perpendicular to forward movement during an individual stride. Positive values indicate movement to the outside |
| Lateral Step Variability | cm | When considering three consecutive foot placements made by the same foot, this describes the variability of perpendicular deviations of the middle foot placement from the line connecting the first and the third. Positive values indicate movement to the outside |
| Toe Off Angle | degrees | The angle of the foot as it leaves the floor at push-off |
| Heel Strike Angle | degrees | The angle of the foot at the point of initial contact |
